# COVID-19 patients exhibit less pronounced immune suppression compared with bacterial septic shock patients

**DOI:** 10.1101/2020.04.03.20049080

**Authors:** Matthijs Kox, Tim Frenzel, Jeroen Schouten, Frank L. van de Veerdonk, Hans J.P.M. Koenen, Peter Pickkers, on behalf of the RCI-COVID-19 study group

## Abstract

At the end of March 2020, there were in excess of 800.000 confirmed cases of coronavirus disease 2019 (COVID-19) worldwide. Several reports suggest that, in severe cases, COVID-19 may cause a hyperinflammatory “cytokine storm”. However, unlike SARS-CoV infection, high levels of anti-inflammatory mediators have also been reported in COVID-19 patients. One study reported that 16% of COVID-19 patients who died developed secondary infection, which might indicate an immune-suppressed state. We explored kinetics of mHLA-DR expression, the most widely used marker of innate immune suppression in critically ill patients, in COVID-19 patients admitted to the ICU.

Twenty-four confirmed COVID-19 patients were included, of which 75% was male and 79% had comorbidities. All patients were mechanically ventilated and exhibited large high levels of inflammatory parameters such as CRP and PCT. Although mHLA-DR expression levels in COVID-19 patients were lower than those observed in healthy subjects, the extent of suppression was less pronounced than observed in bacterial septic shock patients. mHLA-DR expression kinetics revealed no change over time. None of the COVID-19 patients developed a secondary infection.

In conclusion, despite a pronounced inflammatory response, mHLA-DR expression kinetics indicate more moderate innate immune suppression in COVID-19 patients compared with bacterial septic shock patients. These data signify that innate immune suppression as a negative feedback mechanism following PAMP-induced inflammation appears less pronounced in COVID-19.

---

Low monocytic (m)HLA-DR expression is the most widely used marker of innate immune suppression in critically ill patients. We recently showed that, in bacterial septic shock patients, low mHLA-DR expression is prevalent and associated with the development of secondary infections [1]. At the end of March 2020, there were in excess of 800.000 confirmed cases of coronavirus disease 2019 (COVID-19) worldwide, of whom more than 12.000 from the Netherlands. Several reports suggest that patients with severe COVID-19 may suffer from a hyperinflammatory “cytokine storm” [2, 3]. However, unlike SARS-CoV infection, high levels of anti-inflammatory mediators (e.g. IL-10 and IL-4) have also been reported in COVID-19 [3]. Although there are few indications that secondary infections are common in COVID-19 patients, one study reported that 16% of COVID-19 patients who died developed secondary infections [4], which might indicate an immune-suppressed state. Herein, we explored mHLA-DR expression kinetics in a cohort of 24 critically ill COVID-19 patients.

Between March 18^th^-27^th^, all COVID-19 patients admitted to our Intensive Care Unit (ICU) were included in this prospective observational study. COVID-19 was confirmed by two positive RT-PCR tests for SARS-CoV-2 in throat swabs and by CT-scan findings. Fourteen patients were transferred from other ICUs. The median ICU length of stay at the time of study inclusion was 3 days. The study was carried out in accordance with the applicable rules concerning the review of research ethics committees and informed consent in the Netherlands. The ethics committee (CMO Arnhem-Nijmegen) board waived the need for informed consent because of the observational nature of the study and the non-invasiveness of blood withdrawal (all patients had an arterial canula in place, so no venapunctures were performed). All patients or legal representatives were informed about the study details and allowed to abstain from participation. Ethylenediaminetetraacetic acid (EDTA)-anticoagulated blood was stored at 4-8 oC until mHLA-DR expression analysis (performed within 2 hours after withdrawal). Expression levels were determined using the Anti-HLA-DR/Anti-Monocyte Quantibrite assay (BD Biosciences, San Jose, USA) on a Navios flow cytometer and software (Beckman Coulter, Brea, USA). Total number of antibodies bound per cell (mAb/cell) were quantified using a standard curve constructed with Quantibrite phycoerythrin beads (BD Biosciences). All other data were extracted from the electronic patient record. For patients who were transferred from other ICUs, patient characteristics were obtained at admission to our ICU. Data were analysed using SPSS Statistics v22 (IBM, Armonk, USA), and Graphpad Prism v8.3.0 (Graphpad Software, La Jolla, USA).

Patient characteristics are listed in Table 1. In line with previous observations [3], the majority of patients was male and many had comorbidities. The median time from onset of COVID-19 symptoms to ICU admission was 11 days. All patients were mechanically ventilated and exhibited increases in inflammatory parameters (Table 1). As of March 27^th^ 2020, two patients died (at 3 and 4 days post-ICU admission, data of only one timepoint of these patients was recorded), and 22 patients were still in the ICU.

**Table 1.**

| **Characteristics** | **All patients (n=24)** |
| --- | --- |
| Age, years | 69 [61-73] |
| Sex |  |
| Female | 6 (25%) |
| Male | 18 (75%) |
| Body mass index, kg/m<sup>2</sup> | 27.5 [24.3-31.1] |
| Any comorbidities | 19 (79%) |
| Diabetes | 7 (29%) |
| Hypertension | 6 (25%) |
| Cardiovascular disease | 7 (29%) |
| Chronic obstructive pulmonary disease | 3 (13%) |
| Malignancy | 10 (42%) |
| Chronic liver disease | 0 (0%) |
| Chronic kidney disease | 1 (4%) |
| Immunocompromised* | 5 (17%) |
| APACHE II | 17 [11-21] |
| Time from illness onset to ICU admission, days | 11 [8-13] |
| **Medication use** |  |
| Norepinephrine use | 20 (83%) |
| Maximum infusion rate in first 24h on ICU, µg/kg/min | 0.11 [0.07-0.21] |
| Corticosteroids | 1 (4%) |
| Remdesivir | 3 (13%) |
| Chloroquine | 19 (79%) |
| Anakinra | 1 (4%) |
| **Symptoms & (laboratory) parameters** |  |
| Heart rate, bpm | 83 [71-112] |
| Mean arterial pressure, mmHg | 77 [72-81] |
| Fluid balance in first 24h on ICU, mL | 1348 [680-1881] |
| Urine output in first 24h on ICU, mL | 1105 [888-1486] |
| Creatinine, µmol/L | 86 [70-133] |
| Dialysis | 0 (0%) |
| Mechanical ventilation (invasive) | 24 (100%) |
| Tidal volume, mL/kg | 5.3 [4.4-6.0] |
| Respiratory rate, bpm | 21 [20-24] |
| PEEP, cm H<sub>2</sub>O | 12 [10-14] |
| FiO<sub>2</sub>, % | 50 [41-60] |
| P/F ratio | 164 [136-189] |
| 100-200 | 20 (83%) |
| 200-300 | 4 (17%) |
| Thrombocytes, 10<sup>9</sup>/L | 239 [151-274] |
| Leukocytes, 10<sup>9</sup>/L | 8.2 [5.3-11.6] |
| C-reactive protein, mg/L | 301 [157-316] |
| Procalcitonin, µg/L | 0.72 [0.29-3.66] |
| Ferritin, µg/L | 1216 [488-1834] |
| Lactate (highest over last 24h), mmol/L | 1.2 [1.1-1.7] |
| D-dimer, ng/mL | 3075 [1780-4598] |
| Troponin I, ng/L | 23 [13-44] |
| Albumin, g/L | 20 [17-22] |
| Alanine aminotransferase, U/L | 34 [21-41] |
| Aspartate aminotransferase, U/L | 48 [31-73] |
| Creatinine kinase, U/L | 136 [56-357] |
| Lactate dehydrogenase, U/L | 398 [303-499] |
| **Outcome parameters** |  |
| Secondary infections | 0 (0%) |
| Death | 2 (8%) |

Data were obtained at study inclusion and are presented as n (%) or median [IQR]. \*Chronic use of immunosuppressive medication.

Although mHLA-DR expression levels in COVID-19 patients were lower than those observed in healthy subjects (15000-45000 mAb/cell [5]), the extent of suppression was less pronounced than observed in bacterial septic shock patients (geometric mean [95%CI] of 11860 [11035-12746] vs. 5211 [4904-5537] mAb/cell, respectively, p<0.0001, Figure 1A, sepsis data from [1]). mHLA-DR expression kinetics revealed no change over time (Figure 1B). Circulating C-reactive protein concentrations declined over time (Figure 1C), whereas no significant changes in circulating procalcitonin, leukocytes, or ferritin levels were observed (Figure 1D-F). None of the patients developed a secondary infection.

**Figure 1.**
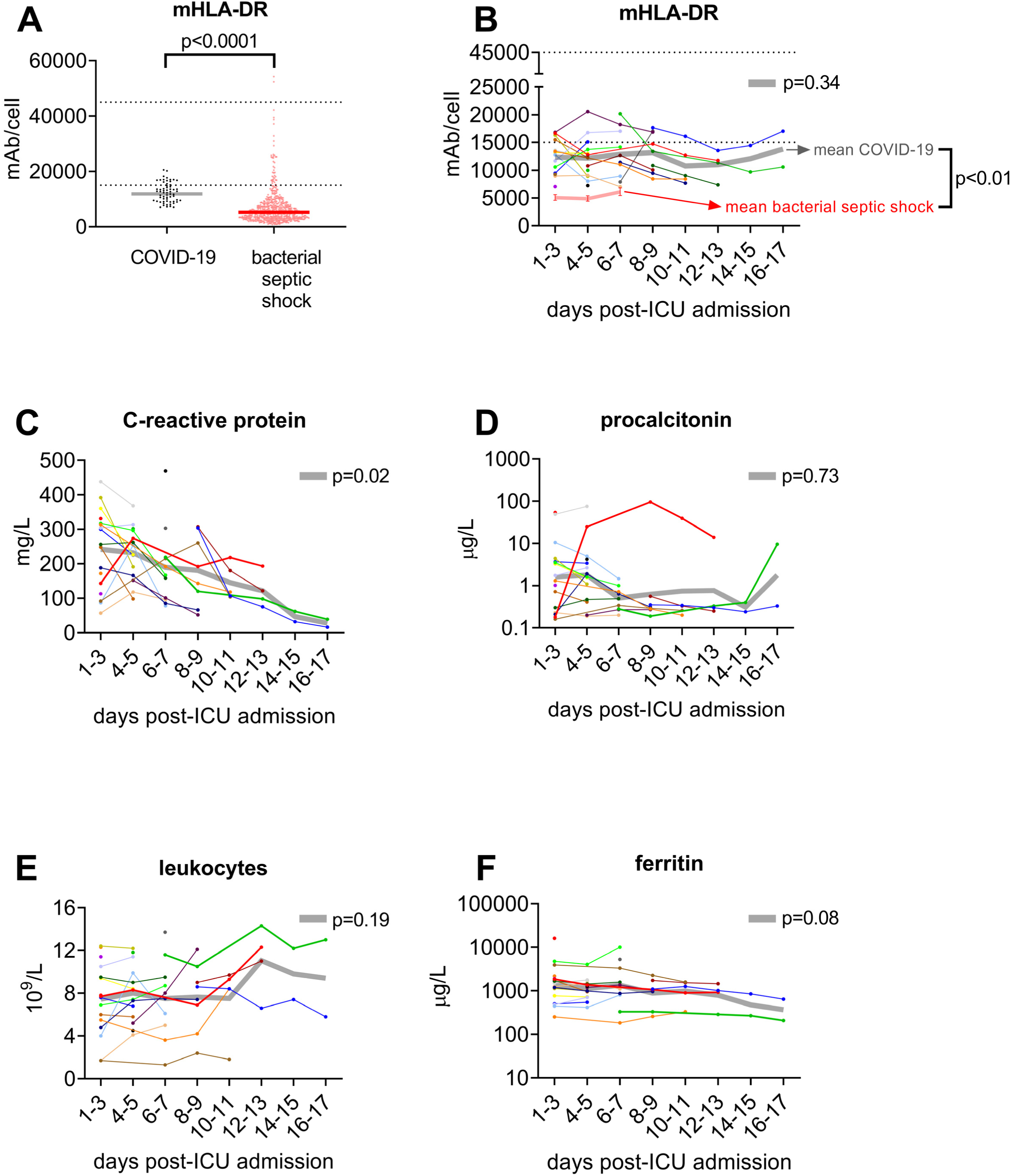
**(A)** mHLA-DR expression in patients with COVID-19 (n=24, multiple time-points) and bacterial septic shock (n=241, days 1-2, 3-4, and/or 6-8 after onset of septic shock, obtained using the same methodology, data recently published [1]). Horizontal line indicates geometric mean. The dotted lines indicate the reference range in healthy subjects [5]. p-value calculated using unpaired t-test on log-transformed data. **(B-F)** Kinetics of mHLA-DR expression, circulating C-reactive protein, procalcitonin, leukocyte numbers, and ferritin in COVID-19 patients (individual data are shown, n=24). The transparent grey line represents mean (panels B, C, and E) or geometric mean (panels C and E) values of the entire cohort. The transparent pink line in panel B represents data obtained from [1] (geometric mean ± 95% CI, please note that values obtained at days 1-2 (n=203), 3-4 (n=205), and 6-8 (n=133) after onset of septic shock are plotted at day 1-3, 4-5, and 6-7, respectively). The dotted lines in panel B indicate the reference range in healthy subjects [5]. p-values next to the transparent grey line represent changes over time in COVID-19 patients, calculated using mixed model analysis (on log transformed data for panels D and F). Differences between COVID-19 and sepsis patients in panel B were analysed using unpaired t-tests on log-transformed data (p<0.0001 on days 1-3 and 4-5, and p=0.0015 on days 6-7).

In conclusion, despite a pronounced inflammatory response in COVID-19 patients, our preliminary results indicate more moderate innate immune suppression compared with bacterial septic shock patients. These findings are in accordance with a low incidence of secondary infections in COVID-19 patients. Therefore, innate immune suppression as a negative feedback mechanism following pathogen-associated molecular pattern-induced inflammation appears less pronounced in COVID-19.

## Data Availability

All data generated or analysed during this study are included in the article.

## Declarations

### Ethics approval and consent to participate

The study was carried out in accordance with the applicable rules concerning the review of research ethics committees and informed consent in the Netherlands. The ethics committee (CMO Arnhem-Nijmegen) board waived the need for informed consent because of the observational nature of the study and the non-invasiveness of blood withdrawal (all patients had an arterial canula in place, so no venapunctures were performed). All patients or legal representatives were informed about the study details and could abstain from participation.

### Consent for publication

Not applicable.

### Availability of data and materials

All data generated or analysed during this study are included in this published article.

### Author contributions

MK and PP designed the study. TF, JS, and FvdV were responsible for data collection. HK performed the flow cytometric analysis. MK performed the statistical analysis and drafted the manuscript. TF, JS, FvdV, HK, and PP critically revised the manuscript. All authors read and approved the final manuscript.

## Acknowledgements

Next to the authors of this letter, the RCI-COVID-19 study group consists of Pleun Hemelaar, Remi Beunders, Johannes van der Hoeven, Sjef van der Velde, Hetty van der Eng, Noortje Rovers, Margreet Klop-Riehl, Jelle Gerretsen, Emma Kooistra, Nicole Waalders, Wout Claassen, Hidde Heesakkers, Tirsa van Schaik, Mihai Netea, Leo Joosten, Nico Janssen, Inge Grondman, Aline de Nooijer, Quirijn de Mast, Martin Jaeger, Ilse Kouijzer, Helga Dijkstra, Heidi Lemmers, Reinout van Crevel, Josephine van de Maat, Gerine Nijman, Simone Moorlag, Esther Taks, Priya Debisarun, Heiman Wertheim, Joost Hopman, Janette Rahamat-Langendoen, Chantal Bleeker-Rovers, Esther Fasse, Esther van Rijssen, Manon Kolkman, Bram van Cranenbroek, Ruben Smeets, Irma Joosten. All of these authors are affiliated to the Radboud Center of Infectious Diseases.

## References (max 6)

1. Leijte GP, Rimmele T, Kox M, Bruse N, Monard C, Gossez M, Monneret G, Pickkers P, Venet F: Monocytic HLA-DR expression kinetics in septic shock patients with different pathogens, sites of infection and adverse outcomes. Crit Care 2020, 24(1):110.

2. Mehta P, McAuley DF, Brown M, Sanchez E, Tattersall RS, Manson JJ, Hlh Across Speciality Collaboration UK: COVID-19: consider cytokine storm syndromes and immunosuppression. Lancet 2020.

3. Huang C, Wang Y, Li X, Ren L, Zhao J, Hu Y, Zhang L, Fan G, Xu J, Gu X et al: Clinical features of patients infected with 2019 novel coronavirus in Wuhan, China. Lancet 2020, 395(10223):497–506.

4. Ruan Q, Yang K, Wang W, Jiang L, Song J: Clinical predictors of mortality due to COVID-19 based on an analysis of data of 150 patients from Wuhan, China. Intensive Care Med 2020.

5. Remy S, Gossez M, Belot A, Hayman J, Portefaix A, Venet F, Javouhey E, Monneret G: Massive increase in monocyte HLA-DR expression can be used to discriminate between septic shock and hemophagocytic lymphohistiocytosis-induced shock. Crit Care 2018, 22(1):213.

